# Making the incomparable comparable: Calibrating patient prioritisation tools for independent surgical procedures onto a common scale

**DOI:** 10.64898/2026.07.23.26358834

**Authors:** Jack Powers, Franz Ombler, Paul Hansen, Ratna Aseervatham, David Grieve, Suzanne Ryan, James M. McGree, Paul Corry

## Abstract

Long waiting times for elective surgery remain a persistent challenge in public health systems. Patient prioritisation tools (PPTs) aim to support equitable decision-making by scoring patients based on clinical and non-clinical factors. However, these tools are often developed independently for specific procedures, resulting in non-comparable scores that hinder consistent prioritisation and risk violating the principles of horizontal equity (equal treatment for equal need) and vertical equity (greater need receives higher priority). This study introduces and demonstrates a calibration method to align multiple procedure-specific PPTs onto a unified severity scale. As a proof-of-concept, ten independently developed PPTs from a general surgery unit at a single Australian public hospital were calibrated using an interactive binary search algorithm to identify clinically equivalent patient states across tools, in collaboration with clinicians from the same unit. Each PPT was then aligned to a common reference procedure, and min-max normalisation was applied to standardise scores onto a shared scale. Internal consistency was assessed by comparing calibration outputs against severity rankings from the same clinical team. The calibrated scores demonstrated strong agreement with the team’s severity judgements (Kendall’s *τ* = −0.734, *p* < 0.001), indicating that the calibration method preserved the clinicians’ intuitive severity rankings. The calibrated scores also preserved the assigned urgency ordering across a cohort of 845 patients. When embedded in an existing dynamic priority formula in simulation, they redistributed waiting time between urgency categories without measurably altering severity concordance. This indicates that the operational effect of the comparable scores depends on the prioritisation formula in which they are embedded, rather than on the calibration itself. These results suggest that the proposed calibration method offers a practical and potentially scalable approach for aligning PPTs across procedures, supporting valid comparisons and enabling more consistent, transparent, and equitable prioritisation. By establishing a common severity scale, this approach addresses a real-world challenge in surgical waiting list management: enabling procedures of inherently different clinical impact to be prioritised fairly and proportionately when competing for shared resources.

## 1 Background

Long waiting times for elective surgery are a common challenge in countries with publicly funded health systems, often resulting in adverse patient outcomes such as deteriorating health and psychological distress [1, 2]. In an effort to reduce these delays and prioritise care more effectively, considerable research has focused on developing patient prioritisation tools (PPTs), which are decision-support systems that assist clinicians to determine the priority of patients and manage waiting lists [3]. These tools aim to reduce average patient waiting times by making waiting lists more equitable, efficient, and transparent by applying standardised and consistent prioritisation methods [3, 4].

In practice, hospital resources do not cater exclusively to a single procedure, condition or specialty, and multiple surgical disciplines must share common resources [5]. Access to these resources is determined by where each patient sits in the queue relative to others, both within their own specialty and across others, as procedures compete for shared capacity. This competition becomes more problematic when prioritisation decisions are made using different tools, which may introduce inconsistencies, inefficiencies, and inequities, often leading to inequitable allocation of resources, long waiting times, and increased morbidity [1, 2, 3, 4, 6, 7, 8, 9].

PPTs use a range of stakeholder-endorsed clinical and non-clinical criteria to reflect a patient’s clinical urgency and capacity to benefit [3, 4, 10]. These criteria are typically structured to support transparent, objective, and standardised prioritisation decisions. PPTs can broadly be categorised as general or specific tools. General PPTs span a wide array of procedures and surgical disciplines using generic criteria to maximise applicability [11, 12, 13, 14, 15, 16]. However, the subjective and less evidence-based nature of these broad criteria often limits their clinical precision [10].

In contrast, specific PPTs focus on a single procedure or condition in a surgical specialty, such as cardiology [17], orthopaedics [18], bariatrics [19, 20], ophthalmology [21, 22] and organ transplantation [23, 24, 25], among others. These tools incorporate criteria that are more explicit, objective, and closely related to condition-specific outcomes [10]. Though they improve prioritisation accuracy within a single procedure, they do not support cross-procedure comparisons, thereby limiting their usefulness in contexts where prioritisation must occur within or across specialties.

This fragmentation introduces challenges for achieving equity in access to care. Horizontal equity refers to the principle that patients with the same clinical need and capacity to benefit should receive equal priority. Vertical equity, by contrast, holds that patients with greater clinical need should receive higher priority than those with lesser need [26, 27, 28]. For example, two patients with comparable pain, functional limitation, and quality-of-life impact undergoing the same procedure should be treated equally (horizontal equity), whereas a patient with more severe symptoms or risk should be treated sooner than one with milder symptoms (vertical equity). Without a common framework to align PPT scores across procedures, both forms of equity are undermined.

The challenge of aligning multiple prioritisation tools relates to the broader literature on multi-criteria decision analysis (MCDA) in healthcare, where structured methods are used to weight and aggregate diverse criteria [29, 30]. However, existing MCDA frameworks typically assume a single decision context; the problem of calibrating scores across independently developed instruments has received limited attention.

General PPTs promote comparability across procedures but at the expense of clinical specificity, whereas specific PPTs enhance within-procedure consistency, but prevent valid comparisons across conditions [4]. Some surgical conditions are inherently more severe than others, making it essential to have a unified scale to ensure that patients from different pathways can be prioritised fairly. For example, a clinical urgency score of 80% in one PPT might not correspond to the same clinical urgency score of 80% in another PPT, and equal scores may not reflect equal urgency. From a measurement perspective, independently developed instruments may exhibit scale parameter heterogeneity, where identical underlying preferences produce different score magnitudes due to differences in error variance and elicitation context [31]. Calibration addresses this heterogeneity by establishing equivalence mappings between scales.

To address this incomparability problem, it is necessary to calibrate the scoring scales of specific PPTs relative to each other. Calibration allows scores from different tools to be mapped onto a common scale, ensuring comparability and interpretability. An example of two calibrated PPTs is illustrated in Figure 1, where calibration establishes linear transformations between their scales so that any point on one scale can be mapped to a point of equal priority on another scale, thereby producing a unified scale.

**Figure 1:**
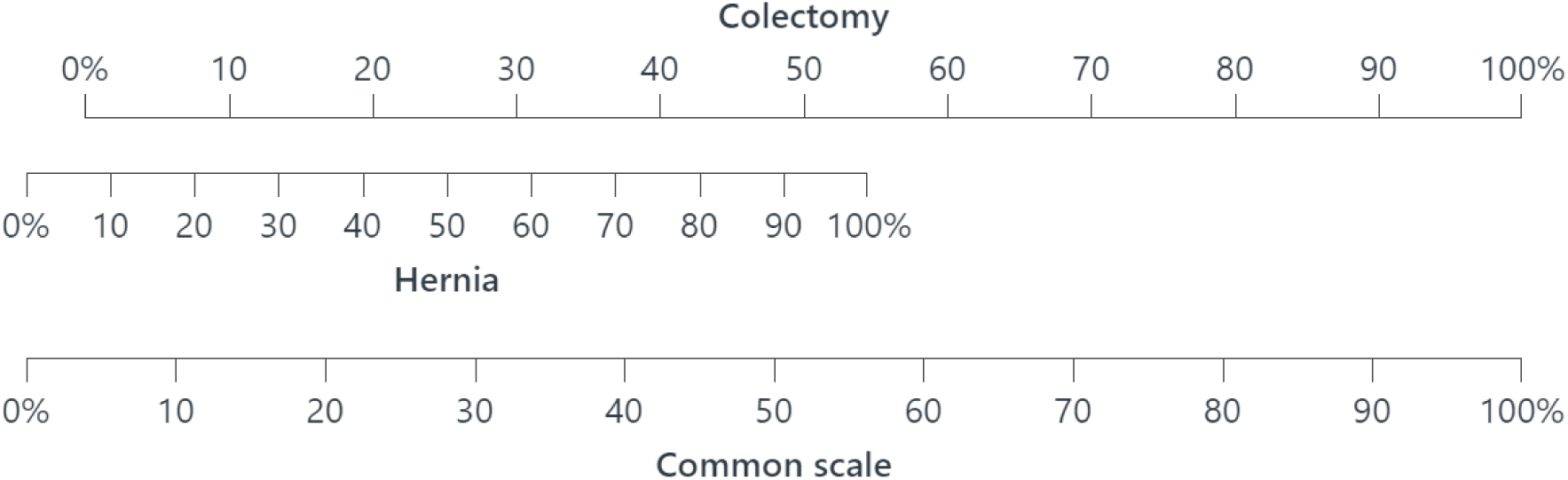
The result of calibrating the colectomy and hernia PPTs, as presented in 1000minds.

This trade-off between comparability and clinical precision is illustrated in Figure 2, which conceptually maps general and specific tools according to their ability to support horizontal and vertical equity. Calibrated specific PPTs enable valid comparisons between different procedures while preserving condition-specific detail, supporting both horizontal and vertical equity.

**Figure 2:**
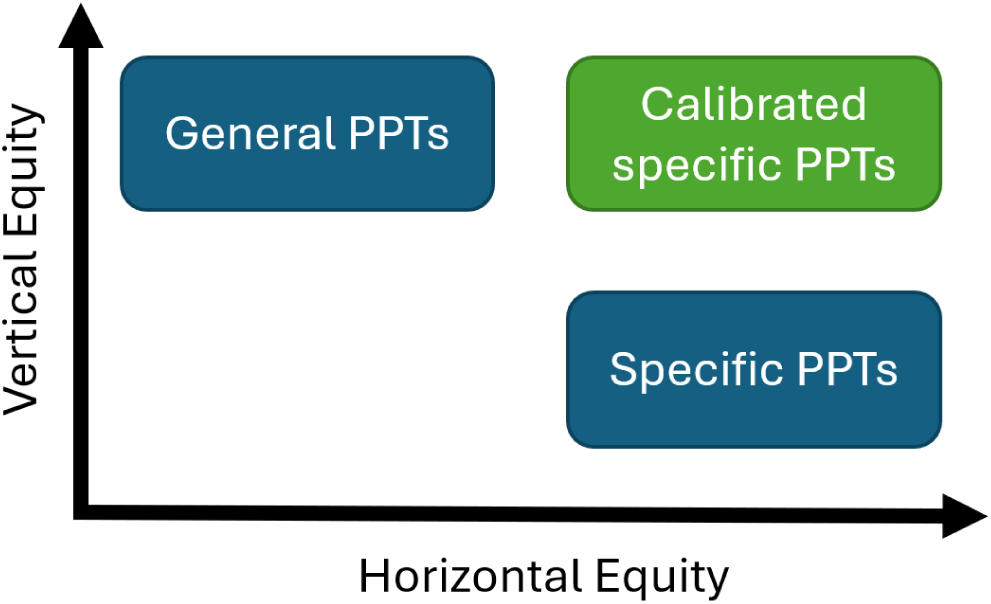
Conceptual positioning of general, specific, and calibrated specific PPTs with respect to horizontal and vertical equity.

Despite its significance, methods for performing such calibrations have been identified as a gap in the literature [4, 32]. A recent systematic review on patient prioritisation methods by Rathnayake et al. [4, p. 14] highlighted that *“none of the studies investigated vertical equity approaches of prioritising methods to re-order the waiting list for different elective surgeries using different weights across disciplines”*.

This study introduces a calibration method that uses an interactive binary search algorithm to rescale procedure-specific PPTs onto a unified, interpretable scale. Unlike previous weighting or calibration methods which do not address vertical equity across different tools, this approach explicitly supports both horizontal and vertical equity and offers a transparent, scalable solution applicable across disciplines. As a proof-of-concept, the method was applied to 10 PPTs in the general surgery specialty, demonstrating the method’s feasibility and broader potential across diverse procedures, clinical contexts, and surgical specialties.

## 2 Methodology

### 2.1 Patient Prioritisation Tool Dataset

The 10 PPTs used in this study were originally developed by a team of general surgeons from a single public hospital in Queensland, Australia, and made publicly available in an earlier simulation study by Powers et al. [33, 34]. The same clinical team participated in both the original PPT development and the subsequent calibration and ranking exercises described in this study, ensuring consistency in clinical judgement throughout but limiting the assessment to internal consistency rather than external validation. Each PPT was created to reflect clinical decision-making for selected surgical procedures within the general surgery specialty (as defined by Royal Australasian College of Surgeons [35]) using criteria and weights informed by clinician input and a structured elicitation method outlined below. These tools produce a clinical factor score between 0 and 1 based on the combination of selected clinical criteria. An example of a PPT for colectomy is shown in Figure 3.

**Figure 3:**
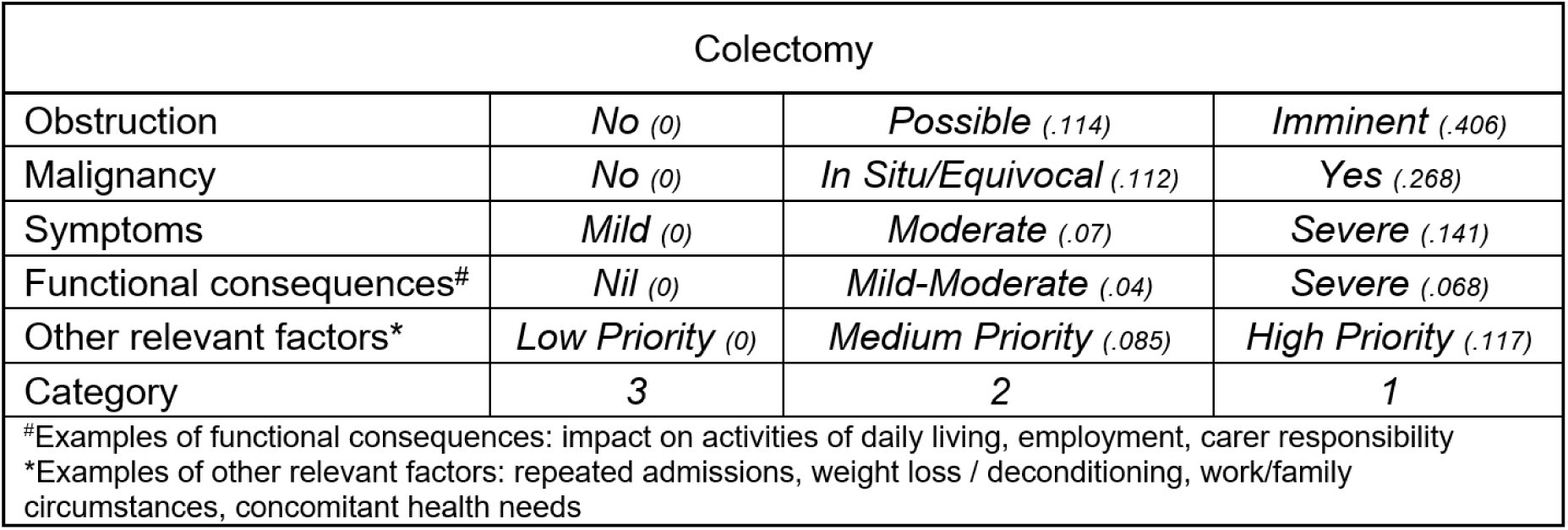
Example of the PPT for the colectomy procedure (with criterion weights in parentheses, [33]

Each time clinicians select the patient state they consider to be of greater clinical urgency, they implicitly reveal the relative importance (or weight) of the criteria and their respective levels, which is then codified by the software. After answering trade-off questions with two criteria, the clinicians answered questions with three criteria (of greater cognitive complexity), thereby maximising the validity and reliability of the derived weights and enabling the PPT patient scores to be treated as interval-scale data appropriate for cross-procedure comparisons. An example of a trade-off question (three criteria) is shown in Figure 5.

The weights for each PPT’s criteria and associated levels were determined using the PAPRIKA method [36], implemented by 1000minds software [37], which simplifies complex multi-criteria decision-making into a series of pairwise comparisons based on expert judgements. The method involved the participating clinicians being repeatedly asked to rank pairs of hypothetical patient health states according to which one has the higher priority for treatment. The pairs of patient health states were described initially on two criteria and later on three criteria at a time, always involving a trade-off between the criteria. An example of a trade-off question (two criteria) is shown in Figure 4.

**Figure 4:**
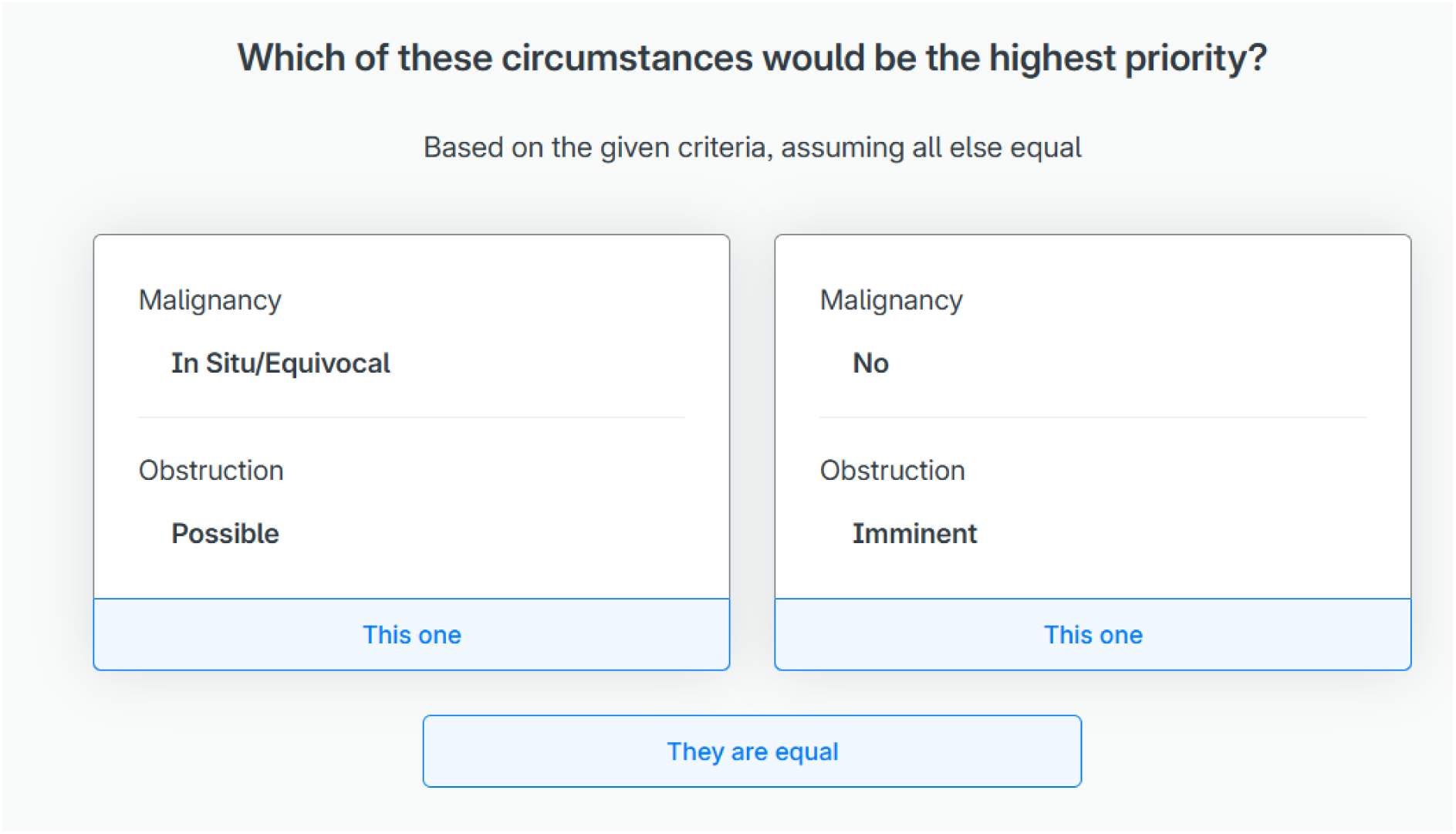
Example of a trade-off for the colectomy procedure as presented in 1000minds.

Although these PPTs can differentiate between patients within the same procedure, supporting horizontal equity within each procedure, the scores from one tool cannot be compared with scores from other tools, a limitation for vertical equity across procedures. To establish comparability, the PPTs need to be calibrated onto a common scale. The calibration method explained below can also be applied to PPTs whose weights are determined using methods other than PAPRIKA.

### 2.2 Calibration Method

#### 2.2.1 Summary

This section outlines the method to calibrate procedure-specific PPTs onto a common scale, enabling fair and consistent comparison of patients across different surgical procedures. The calibration involved a structured, multi-step process: a clinician ranking exercise to independently assess the severity of extreme patient states, then an interactive binary search algorithm in which clinicians were presented with structured pairwise comparisons between hypothetical patient states from different PPTs to identify points of equal severity across scales, allowing scores from inherently different procedures to be transformed onto a unified interval-scale measurement, and a final validation step to confirm the alignment between the calibrated scores and clinical judgement.

#### 2.2.2 Clinical Severity Ranking of Procedures

To assess internal consistency, the same group of general surgeons who developed the original PPTs participated in an in-person consensus ranking exercise to evaluate the most and least severe patient states for each of the 10 procedures. These states were defined as the highest and lowest possible scores within each PPT (i.e. severity scores of 1 and 0, respectively), representing the clinical extremes. Participants were repeatedly presented with pairwise comparisons between hypothetical patient scenarios and asked to rank them based on clinical urgency. The resulting ranked list reflected clinicians’ intuitive understanding of relative severity across procedures and served two purposes: it provided an independent benchmark for validating the calibrated scores, and it offered insight into the severity span of the procedures, aiding in the selection of a suitable base PPT (i.e., a common comparison against every other PPT). Colectomy was ultimately chosen as the base procedure for calibration, based on the ranking exercise, as both clinical extremes were familiar to the clinicians and generally encompassed the other procedures on the ranked severity spectrum.

#### 2.2.3 Scale Transformation Principles

A calibration method was developed to calibrate the PPTs onto a common scale, implemented through calibration surveys using 1000minds software (www.1000minds.com) [37]. The surveys are publicly accessible via 1000minds and provided in Appendix A.

In the simplest case – when both PPTs being compared have interval-scale measurement properties [38] – only two equivalence mappings between the scales are required to define a transformation function. This function can either convert one scale into another or establish a unified scale spanning both tools. As illustrated in Figure 1, two points of equivalence are sufficient to align two interval scales.

A familiar analogue is the transformation between Fahrenheit (F) and Celsius (C) temperature scales: knowing that 32^∘^F = 0°C and 212^∘^F = 100°C (or any other two mappings) enables the derivation of the linear transformation 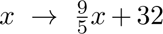. This concept of linear transformation can be extended to PPT calibration: when the scales have interval properties, two equivalence points are sufficient to determine a transformation that preserves clinical meaning across the scales. In such cases, a constant difference (denoted as Δ) in score represents a consistent difference in clinical severity across the scale, unlike ordinal scales, which preserve only rank order.

For PPTs developed using 1000minds software, interval-scale measurement can be inferred from the software’s reported accuracy statistic and by examining the indicated possible range of weights; if higher accuracy is required, this can usually be achieved by completing trade-offs involving three or more criteria, as illustrated in Figure 5 (in contrast to the trade-off question presented in Figure 4) [38].

**Figure 5:**
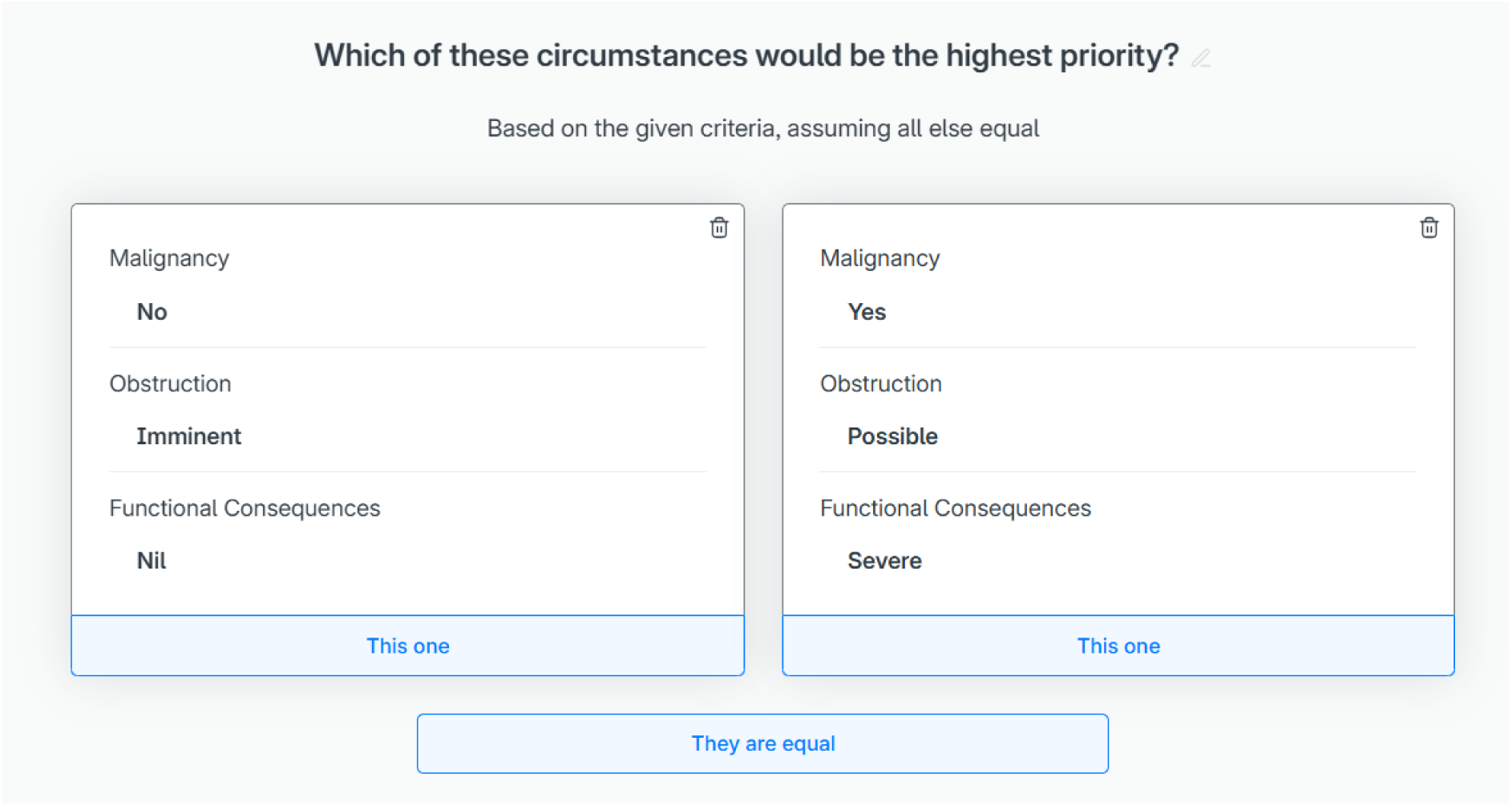
Example of a three-level trade-off for the colectomy procedure, as presented in 1000minds.

To clarify the operational meaning of this assumption: a PPT possesses interval-scale measurement properties when equal numerical differences in its scores correspond to equal differences in clinical priority, regardless of where on the scale those differences occur. Specifically, the difference in severity between scores of 0.2 and 0.4 should carry the same clinical meaning as the difference between 0.6 and 0.8. This property distinguishes interval scales from merely ordinal scales, which preserve only the rank ordering of patients without guaranteeing that score differences are commensurable.

The PAPRIKA method establishes interval-scale properties through its elicitation procedure [36, 38]. As criteria weights are derived from a complete set of pairwise trade-off comparisons, the resulting scores reflect the relative magnitude (not merely the direction) of clinician preferences. The accuracy statistic reported by 1000minds quantifies the degree of transitive consistency across these pairwise judgements: if a clinician indicates that state ***A*** is more severe than state ***B***, and ***B*** more severe than ***C***, then transitive consistency requires that ***A*** is also more severe than ***C***. A high accuracy value indicates that the elicited weights faithfully represent a coherent interval scale, whereas low accuracy would suggest that the preference ordering contains circular inconsistencies incompatible with interval measurement. This assumption is critical to the calibration approach: the linear transformation between two PPT scales (analogous to the Fahrenheit–Celsius conversion above) is valid precisely because both scales possess interval properties. If the scales were only ordinal, a linear mapping would not preserve clinical meaning, and a more complex, potentially non-parametric, transformation would be required.

To establish equivalence mappings between PPTs, an interactive binary search (or bisection) algorithm was employed to identify where a reference point on one scale corresponds to a point on another scale. The reference points used were the most and least severe states within each PPT. Binary search was chosen for its computational efficiency and conceptual simplicity, enabling rapid convergence with minimal comparisons. Unlike exhaustive search methods, which may require evaluating many possible health states, binary search iteratively halves the search space, thereby reducing cognitive load for participants. Furthermore, it supports a transparent and reproducible decision process, with each step traceable and reviewable.

As detailed in Sullivan et al. [39], this particular implementation of binary search differs from standard implementations in two important ways, with the objective of reducing the cognitive burden on participants. Firstly, the algorithm begins at the top of the scale instead of the midpoint. This starting point ensures that if a participant selects the base PPT as more severe, they are only required to make a single comparison. Secondly, the binary search does not evaluate all possible states representable by the PPT. For example, for a PPT with six criteria and four levels each, there are 4^6^ = 4096 possible combinations. To better manage and reduce the number of potential states, a subset of these states is formed by rounding their values (clinical factor scores) to two decimal places, grouping them by value and selecting one state from each grouping. This yields a subset with a maximum of 101 states, where each state has a value between zero and one rounded to two decimal places.

Participants (i.e. clinicians), either individually or working as a group, were initially presented with a choice between two hypothetical patients representing the most severe states on each of the two PPTs under comparison. For example, in the comparison between the hernia and colectomy PPTs shown in Figure 6, the PPT on the right-hand side served as the “base” procedure, anchoring the calibration. As the method progressed, the health states presented on the left were updated adaptively based on the participants’ previous responses.

**Figure 6:**
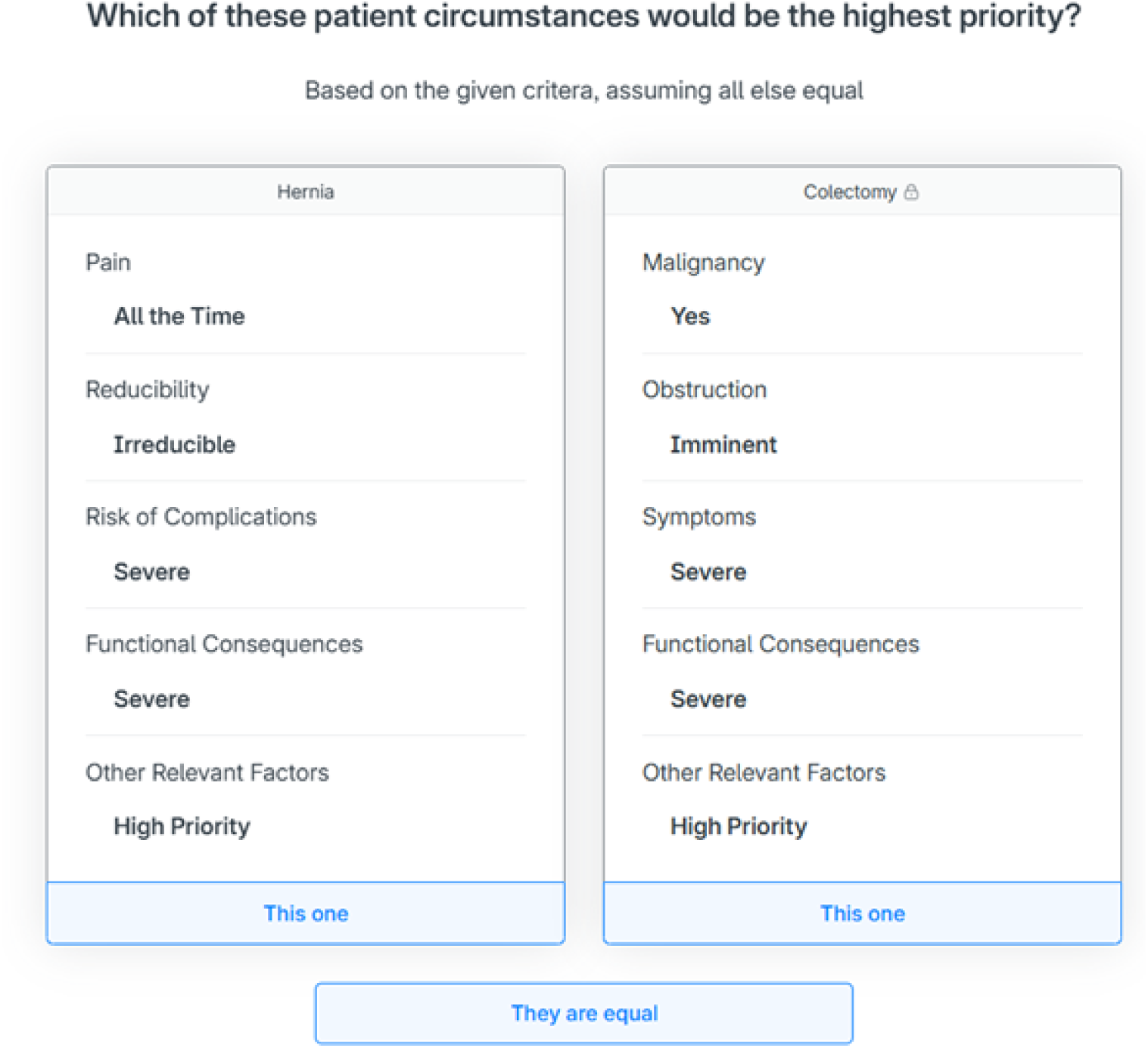
Example of the first comparison of the calibration procedure (as portrayed in Figure 8a) for the conditions of hernia and colectomy, as presented in 1000minds.

The notation *α*_*b*_ is used to denote the set of criteria presented in PPT *α* that results in a factor score of *b* (or as close to *b* as the weights allow), where *b* is a value between zero and one. Using this notation, the PPTs presented in Figure 6 can be denoted as Hernia_1_ and Colectomy_1_, indicating the most severe states of the hernia and colectomy PPTs respectively.

The following subsections detail key elements of the calibration method, including mechanisms for changing the base PPT and handling potential contradictions in clinician responses. A complete flowchart of the calibration process is provided in Figure 7.

**Figure 7:**
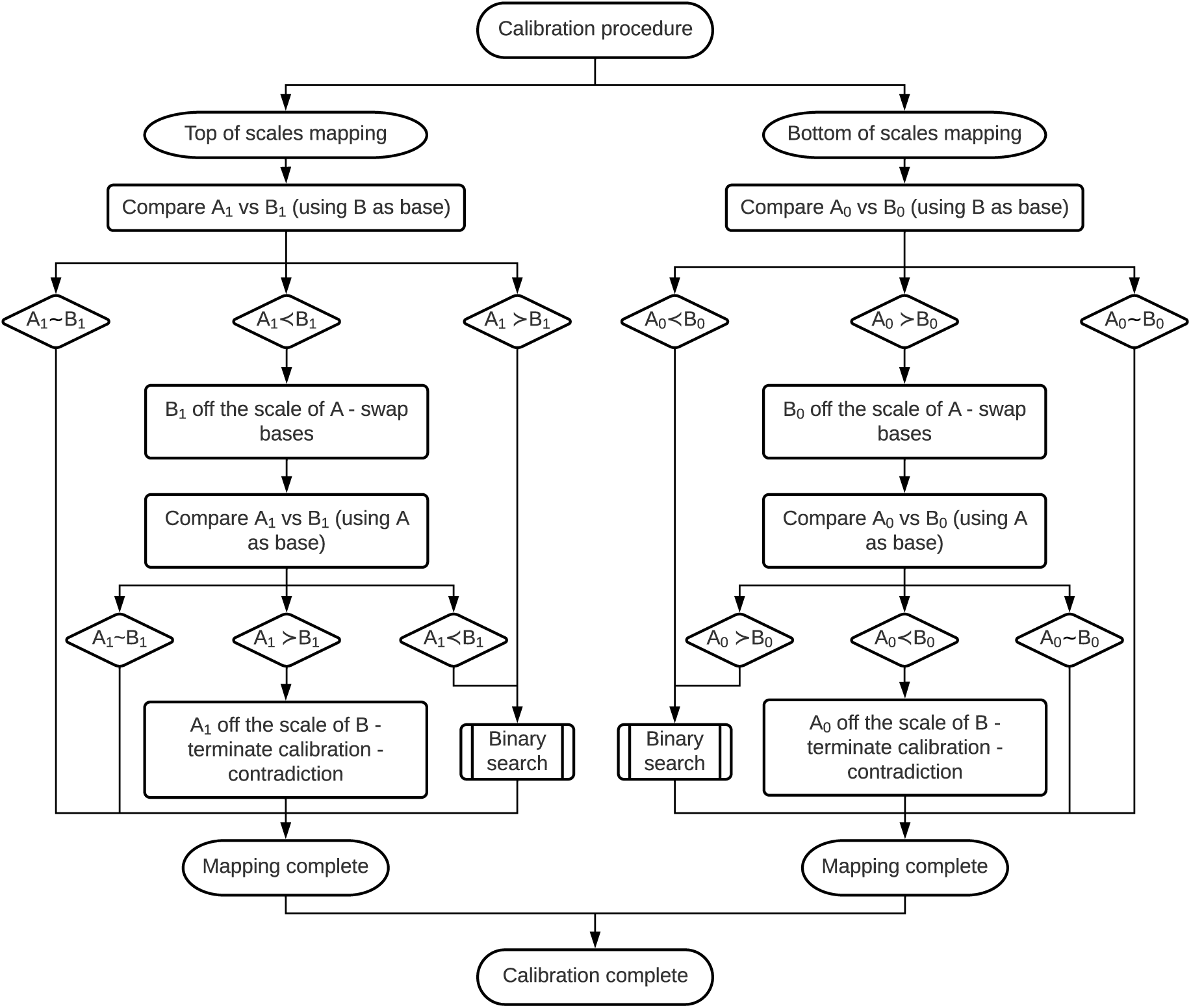
Calibration procedure methodology.

#### 2.2.4 Top-of-Scales Mapping

The algorithm begins by presenting the participant with a choice between the most severe states of both PPTs, ***A***_1_ *and* ***B***_1_. If the participant selects ***A***_1_ ≻ ***B***_1_, the algorithm bisects the scale of ***A*** and presents the next choice of ***A***_0.5_ *vs* ***B***_1_. Depending on the participant’s choice, the algorithm repeatedly bisects the scale of ***A*** and presents another health state. For example, if ***A***_0.5_ ≻ ***B***_1_, ***A***_0.25_ *vs* ***B***_1_ is posed next. Conversely, if ***A***_0.5_ ≺ ***B***_1_, then ***A***_0.75_ *vs* ***B***_1_ is posed. This process is repeated, with the scale of ***A*** being continually bisected until the algorithm terminates.

The binary search algorithm terminates if at any stage: the participant deems two states to be equal (i.e. ***A***_…_ ∼ ***B***_1_); enough comparisons have been completed to infer where the scales of each PPT are equal (to a finite precision); or if the participant selects ***A***_1_ ≺ ***B***_1_, in which case the base is swapped (as this choice would place ***B***_1_ off the scale of ***A*** – see Change of Base below) and the binary search restarts. An illustration of the binary search algorithm steps is shown in

Figure 8, where the participant’s choices are indicated. The comparison presented in Figure 6 is representative of the choice portrayed in Figure 8a.

**Figure 8:**
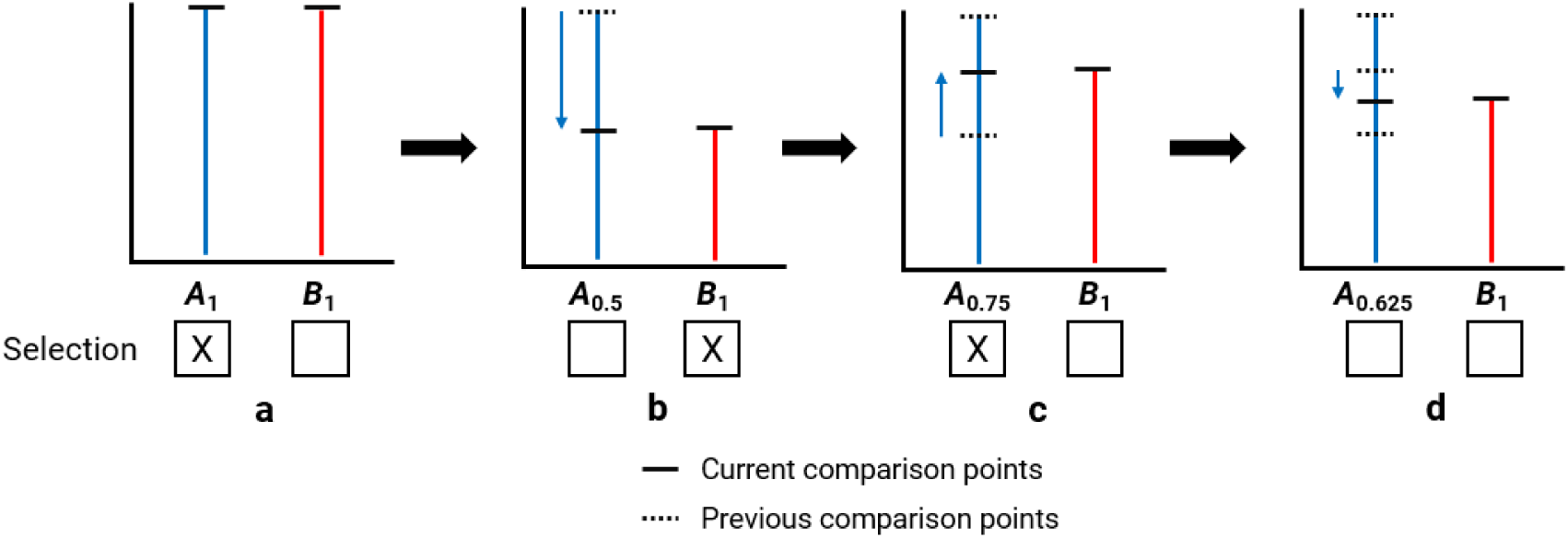
Binary search algorithm (selection is indicative of the choice made that is deemed to be of highest preference, as presented in each sub-figure).

#### 2.2.5 Bottom-of-Scales Mapping

To map the least severe states, the algorithm presents the participant with a choice between ***A***_0_and ***B***_0_. If the participant selects ***A***_0_ ≺ ***B***_0_, the algorithm bisects the scale of ***A*** and presents the next comparison of ***A***_0.5_ vs ***B***_0_, continuing the binary search. If instead the participant selects ***A***_0_ ≻ ***B***_0_, the algorithm terminates and swaps the base PPT, as this selection would place ***B***_0_off the scale of ***A*** (see Change of Base below).

#### 2.2.6 Change of Base

As mentioned above, if a comparison is deemed to be off the scale of the base PPT, the binary search algorithm terminates and the base PPTs are swapped. For example, if ***A***_1_ vs ***B***_1_ is presented to the participant (where PPT B is the current base) and they select ***A***_1_ ≺ ***B***_1_, this choice results in ***B***_1_ being off the scale of ***A***, as shown in Figure 9a.

**Figure 9:**
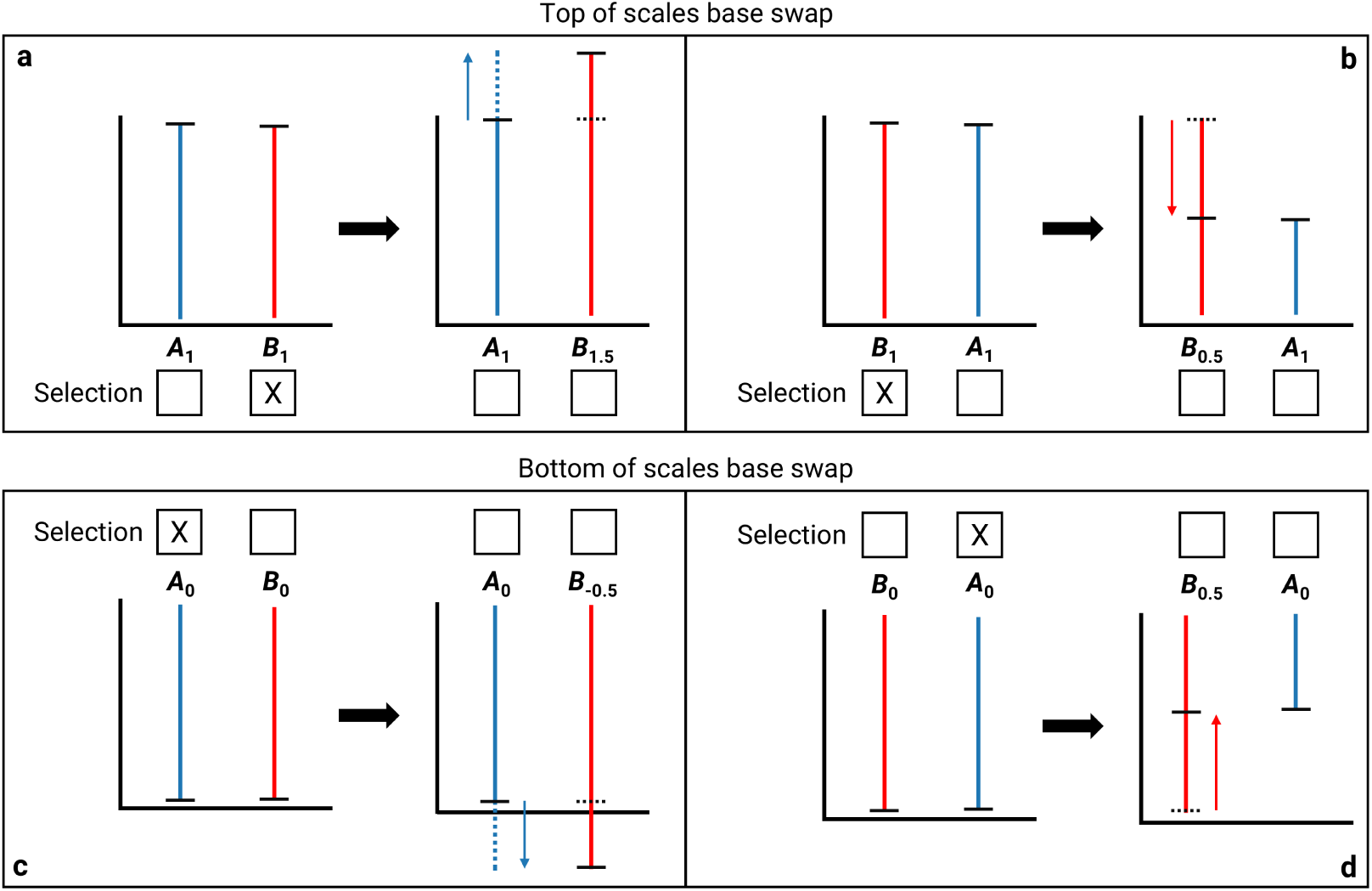
Change of base procedure for top of scales (a & b) and bottom of scales (c & d) (selection is indicative of the choice deemed to be of highest preference, as presented in each sub-figure) procedure, confusion about criteria, or disagreement with the underlying weights of the PPT criteria.

Consequently, to ensure the resultant comparison remains within the bounds of either PPT, the base PPT is swapped and the binary search restarts, as demonstrated in Figure 9b. Similarly, the process of calibrating ***A***_0_vs ***B***_0_ and the subsequent base swap if the participant selects ***A***_0_ ≻ ***B***_0_ can be seen in Figure 9 c & d.

#### 2.2.7 Contradiction

Depending on the participant’s choices, there is a possibility that the calibration method cannot be completed due to contradictory selections. For example, if the participant were presented with ***A***_1_vs ***B***_1_ and selected ***A***_1_ ≺ ***B***_1_ when the base was PPT B, and subsequently selected ***A***_1_ ≻ ***B***_1_ when the base changed to PPT A, the algorithm would terminate due to a contradiction between these comparisons; i.e. PPT A cannot be off the scale of PPT B and vice versa simultaneously.

Although contradictions in participant choices during calibration can lead to early termination of the algorithm, they serve as valuable quality checks on participant engagement, comprehension, and tool validity. A high rate of contradictions may indicate unfamiliarity with a

#### 2.2.8 Application of Calibration Method and Scale Normalisation

The calibration method was conducted via a series of structured surveys completed by a group of clinicians. Each of the nine remaining PPTs was individually calibrated against the base PPT (colectomy), using the outlined binary search process. For each PPT pairing, clinicians were presented with a sequence of pairwise comparisons between patient states to determine points of equal clinical urgency. Each calibration pairing was completed by consensus among the clinician group. As described in Clinical Severity Ranking of Procedures above, colectomy was selected as the base procedure based on its familiarity to the participating clinicians and its encompassing position across the clinician-derived severity rankings, making it a suitable anchor for cross-procedure calibration.

These comparisons yielded a set of transformation functions aligning the scales of each procedure with that of colectomy. To produce a unified scoring system across all procedures, min-max normalisation was applied to the top and bottom transformation values from each calibrated pair. This process scaled all scores to fall within the [0, 1] range, facilitating cross-procedure comparisons on a common interval scale. The min-max normalisation was defined as 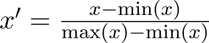 where *x* represents the set of all scale transformations and *x*′ is a set of the normalised scale transformations.

The outcome of this process was a fully calibrated and normalised scale across all 10 general surgery PPTs, enabling valid and interpretable comparisons of patient severity irrespective of the original tool used.

#### 2.2.9 Validation Approach

To assess the validity of the calibrated scale, the final transformed scores for each procedure (Application of Calibration Method and Scale Normalisation) were compared to clinician-derived rankings established in the severity ranking exercise (Clinical Severity Ranking of Procedures). Specifically, the top and bottom calibrated scores for each procedure were matched to their corresponding positions in the consensus ranking.

To evaluate the degree of alignment, the calibrated scores were correlated with the clinicians’ consensus ranking. As these rankings are ordinal and contain tied positions, rank-based association measures were preferred to the Pearson correlation, which is most appropriate for interval-scaled, bivariate-normal data [40]. Kendall’s tau (*τ*) was therefore adopted as the primary measure of agreement, with Spearman’s rank-order correlation (*ρ*) and Pearson’s correlation coefficient (*r*) reported as confirmatory checks. These analyses tested whether the relative ordering of procedures on the unified scale reflected the clinicians’ intuitive judgement of the high- and low-severity states.

### 2.3 Operational Evaluation

To assess the operational impact of calibration, the calibrated cross-procedure scores were used as the clinical-factor term of the dynamic priority score of Powers et al. [33] and evaluated through that study’s discrete-event simulation of the elective surgery waiting list. Both configurations utilised an identical priority formula, 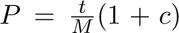, where *P* is the priority score, *t* is the time waited in days, *M* is the maximum recommended waiting time for the patient’s urgency category, and *c* is the clinical-factor score; the only difference between them was whether *c* was the raw procedure-specific score or the calibrated cross-procedure score introduced in this study. As such, any difference in the resulting waiting times is attributable to the calibration alone.

## 3 Results

### 3.1 Procedure Severity Rankings

Table 1 summarises the severity rankings derived from expert clinicians, representing their intuitive assessment of clinical severity across procedures. These rankings provided a baseline for subsequent validation of the calibrated scale.

**Table 1:** Procedure severity ranking (lower values indicate higher preference).

| Procedure | Low | High |
| --- | --- | --- |
| Breast | 12.5 | 6 |
| Cholecystectomy | 17.5 | 4.5 |
| Colectomy | 17.5 | 2 |
| Diagnostic laparoscopy | 14 | 7.5 |
| Hernia | 17.5 | 10 |
| Pancreatic resection | 17.5 | 3 |
| Perianal | 17.5 | 4.5 |
| Reversal of stoma | 12.5 | 9 |
| Skin lesion excision | 11 | 7.5 |
| Thyroidectomy | 17.5 | 1 |

### 3.2 Inter-Procedure Weightings and Calibrated Scale

The calibration procedure successfully aligned each PPT with expert clinicians’ judgements of relative clinical severity. Table 2 summarises the results of calibrating each of the nine general surgery PPTs against the base PPT (colectomy). Each entry denotes an independent application of the calibration procedure and indicates the alignment of the most and least severe states for each procedure relative to colectomy.

**Table 2:** Calibration procedure results (rounded to three decimal points).

| # | Procedure A | Procedure B | Low: B on A | High: B on A |
| --- | --- | --- | --- | --- |
| 1 | Colectomy | Breast | −0.042 | 1.000 |
| 2 | Colectomy | Cholecystectomy | 0.020 | 1.000 |
| 3 | Colectomy | Diagnostic laparoscopy | −0.067 | 0.540 |
| 4 | Colectomy | Hernia | −0.041 | 0.545 |
| 5 | Colectomy | Pancreatic resection | −0.224 | 1.019 |
| 6 | Colectomy | Perianal | −0.032 | 0.770 |
| 7 | Colectomy | Reversal of stoma | −0.013 | 0.495 |
| 8 | Colectomy | Skin lesion excision | −0.052 | 0.555 |
| 9 | Colectomy | Thyroidectomy | −0.098 | 1.528 |

Applying min-max normalisation to these calibrated transformations produced a unified scoring scale suitable for direct inter-procedure comparisons. These normalised scale transformation values are shown in Table 3. The effects of calibration, comparing original and calibrated scales respectively, can be seen in Figure 10a&b.

**Figure 10:**
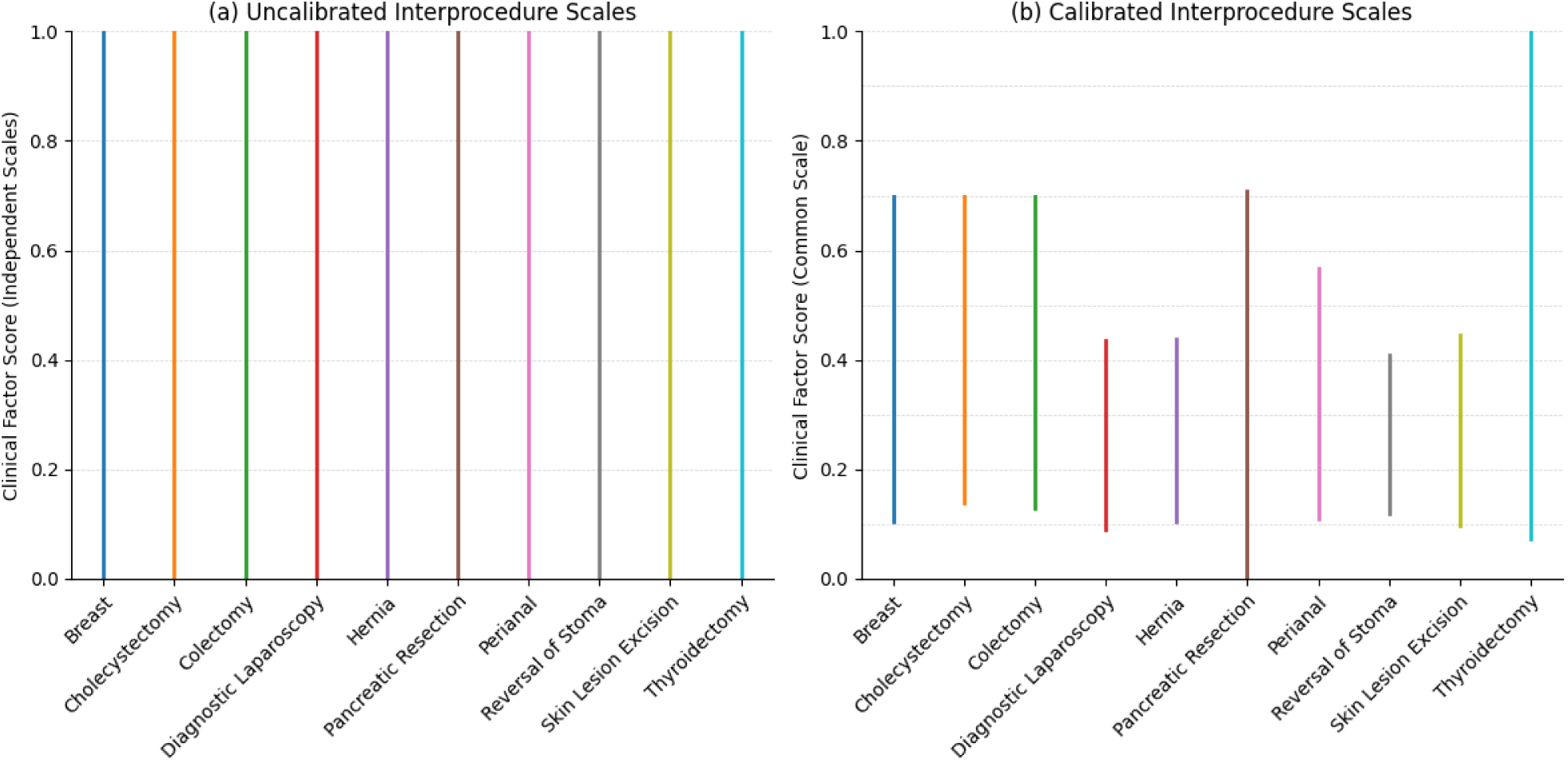
Uncalibrated (a) and calibrated (b) scale transformations.

**Table 3:** Normalised scale transformations (rounded to three decimal points).

| Procedure | Low | High |
| --- | --- | --- |
| Breast | 0.104 | 0.699 |
| Cholecystectomy | 0.139 | 0.699 |
| Colectomy | 0.128 | 0.699 |
| Diagnostic laparoscopy | 0.090 | 0.436 |
| Hernia | 0.104 | 0.439 |
| Pancreatic resection | 0.000 | 0.709 |
| Perianal | 0.110 | 0.567 |
| Reversal of stoma | 0.120 | 0.410 |
| Skin lesion excision | 0.098 | 0.445 |
| Thyroidectomy | 0.072 | 1.000 |

### 3.3 Validation Against Clinician Severity Rankings

The validity of the calibrated scales was evaluated by comparing the final transformed scores to clinician-derived rankings from the severity ranking exercise. Correlation analyses assessed the consistency between the clinicians’ severity rankings and the calibrated scale scores for both the most and least severe states.

The correlation analyses indicated strong, statistically significant agreement between the calibrated scores and clinician severity rankings. The primary measure, Kendall’s *τ* = −0.734 (95% CI [−0.851, −0.547] [40], *p* < 0.001), was in agreement with Spearman’s rank-order correlation *ρ* = −0.873 (*p* < 0.001) and Pearson’s correlation coefficient *r* = −0.919 (*p* < 0.001). The negative sign reflects that the states clinicians judged most severe were assigned the lowest rank numbers while receiving the highest calibrated scores. As such, the calibration preserved the clinicians’ relative severity ordering; establishing broader generalisability would, however, require validation involving clinicians from other institutions.

### 3.4 Consistency with Clinical Urgency Categories

The validation above compares the calibrated scale against clinician rankings of hypothetical extreme states. A complementary test is whether the calibrated scores remain consistent with clinical judgement when applied to a full cohort of real patients, and in particular whether this consistency is maintained *across* procedures, where the uncalibrated procedure-specific scores are not directly comparable. To assess this, the calibration was applied to the 845 patients of the previously published dataset [34], and each patient’s calibrated severity score was compared against the urgency category assigned at the time of listing. As both the urgency category and the underlying clinical-factor score encode clinical severity, a degree of agreement is expected by construction; the informative question is therefore whether the calibrated scale preserves this agreement once scores are rendered comparable between procedures.

The calibrated scores are consistent with the assigned urgency categories (Kendall’s *τ* = −0.635, *p* < 0.001; Spearman’s *ρ* = −0.768, *p* < 0.001; Pearson’s *r* = −0.707, *p* < 0.001), where the negative sign reflects that higher calibrated severity corresponds to a more urgent, lower-numbered category. This consistency is maintained across procedures: among all comparisons between a Category 1 and a Category 3 patient drawn from *different* procedures, the Category 1 patient receives the higher calibrated score in approximately 98% of cases. Only two of the 339 Category 1 patients (0.6%) receive a calibrated score below the median Category 3 patient (one a skin lesion excision and the other a pancreatic resection); both have low clinical-factor scores relative to their assigned urgency and belong to procedures with low calibration floors. This relationship can be seen in Figure 11.

**Figure 11:**
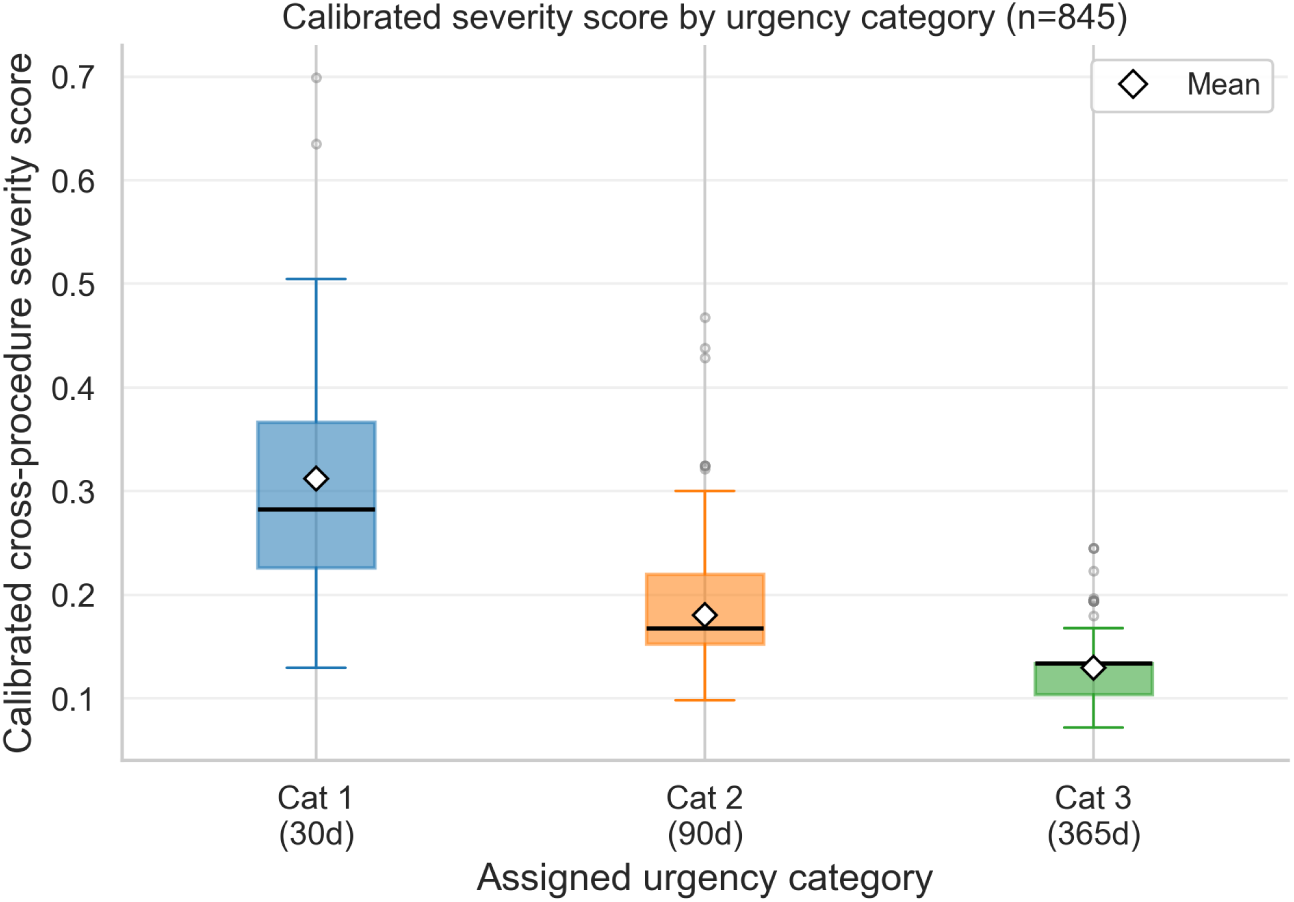
Calibrated cross-procedure severity score by assigned urgency category for the 845-patient cohort. Higher (less urgent) categories correspond to lower calibrated severity, and the separation is maintained across procedures.

For reference, the uncalibrated procedure-specific scores yield a marginally stronger association with the urgency categories (Spearman’s *ρ* = −0.782; cross-procedure concordance 99.2%) than the calibrated scores (*ρ* = −0.768; 98.4%). This small difference reflects the compression of the score range introduced by calibration rather than any loss of clinical validity; within any single procedure the two scales induce an identical ordering. The value of calibration lies not in improving agreement with the existing categories, which the procedure-specific scores already encode, but in rendering scores comparable across procedures while preserving that agreement.

Overall, these results indicate that the calibrated scale preserves the clinically established urgency ordering while rendering severity comparable across procedures. As such, the calibration reconciles within-procedure prioritisation with cross-procedure comparability without inverting the urgency relationships embedded in current clinical practice. This cohort-level agreement should be read as a consistency check rather than independent confirmation, since the urgency category is itself informed by clinical severity; the preservation of the ordering across procedures nonetheless provides support for the calibrated scale beyond the extreme-state rankings reported above.

### 3.5 Operational Impact of Calibration on Waiting Times

The analyses above establish that the calibrated scale is consistent with clinician judgement at the level of the scores themselves, both for hypothetical extreme states and for the assigned urgency categories of a full patient cohort. How the scale behaves once placed in a dynamic prioritisation system and allowed to govern the order in which patients are treated is a separate, operational question, concerning the effect of calibration on waiting times rather than its validity.

In the simulation, Category 3 patients, whose maximum recommended waiting time is the most permissive at 365 days, waited a median of about 14 days less under the calibrated scores (336 to 322 days pooled across replications; the per-replication change averaged −13.6 days, was negative in all twenty replications, and carried a 95% confidence interval of [−16.5, −10.6] days). Category 1 patients waited a median of 3 days more (28 to 31 days), while Category 2 patients were largely unaffected, their pooled median shifting from 84 to 87 days against a mean and compliance that moved by no more than 0.1 days and 0.1 percentage points respectively. These changes redistributed waiting time rather than reducing it overall: treating the longest-waiting Category 3 patients sooner moved some Category 1 patients past their tighter 30-day target, and aggregate compliance with the category targets fell from 61.9% to 56.3%, a fall driven by Category 1, whose patients outnumber Category 3 patients roughly sevenfold, while Category 3 compliance itself rose from 74.3% to 84.3%. The cross-procedure severity concordance, the proportion of patient pairs drawn from different procedures in which the patient with the higher calibrated severity score experiences the shorter wait, is statistically indistinguishable between the two systems (0.75 under each; paired difference −0.001, 95% confidence interval [−0.003, +0.001], *p* = 0.21). Both configurations are evaluated against this same calibrated reference, with the uncalibrated configuration’s raw scores mapped through the published anchors, so the comparison favours neither system; the calibration therefore does not measurably change how well treatment order tracks clinical severity. These per-category statistics can be seen in Table 4, which also reproduces the three-category baseline of the original study [33] for reference, and the distributions in Figure 12.

**Figure 12:**
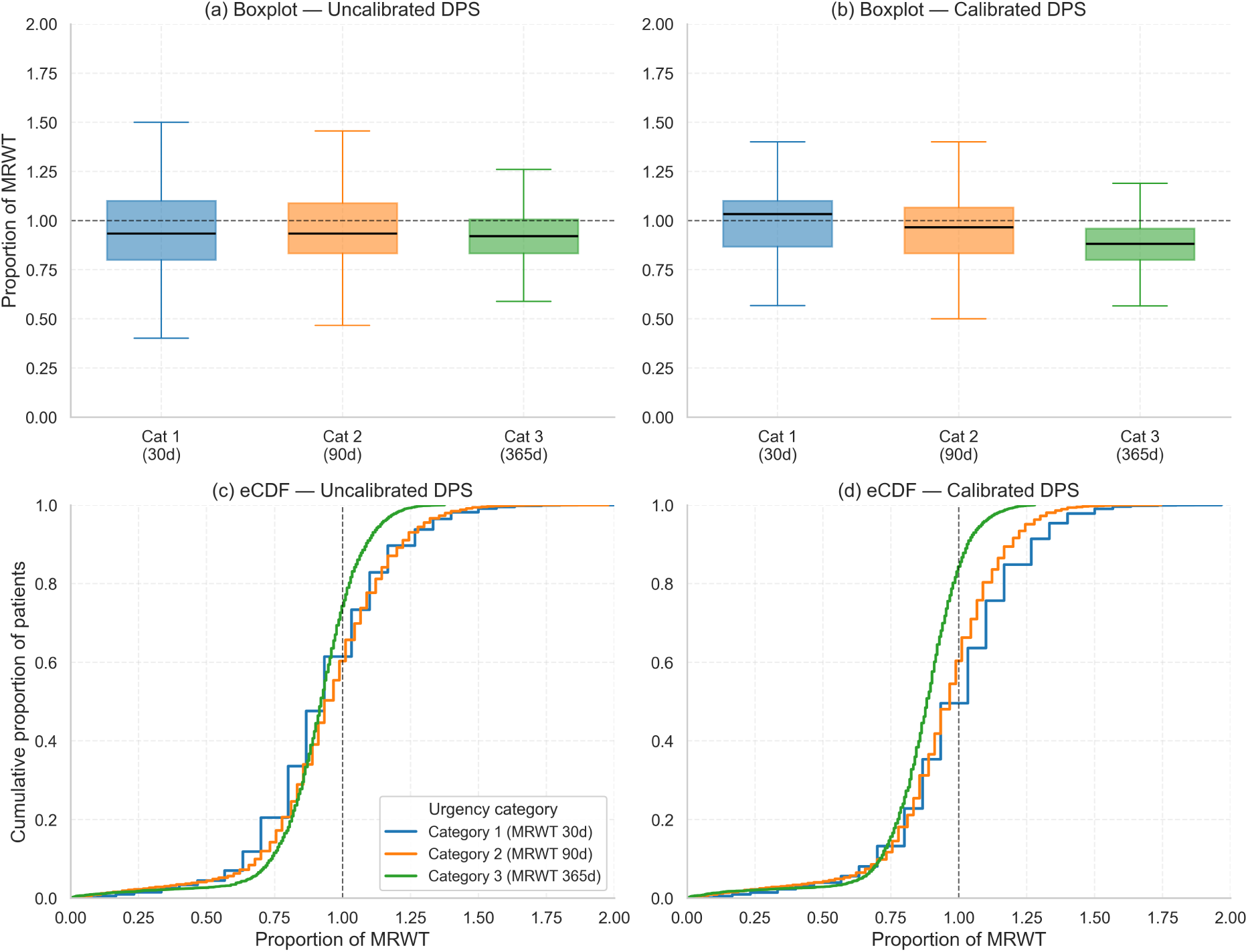
Waiting time as a proportion of the maximum recommended waiting time (MRWT), by urgency category, under the uncalibrated (procedure-specific) Dynamic Priority Score of Powers et al. [33] and its calibrated (cross-procedure) counterpart, across the twenty simulation replications. Panels (a, b) show per-category box plots and panels (c, d) the corresponding empirical cumulative distribution functions, with the dashed line marking the MRWT.

**Table 4:** Summary statistics of patient waiting times in days (proportion of MRWT) under the three-category baseline and the uncalibrated and calibrated Dynamic Priority Score, by urgency category.

| System | Cat | $n$ | Mean | Std | 5% | Q1 | Median | Q3 | 95% | Compliance |
| --- | --- | --- | --- | --- | --- | --- | --- | --- | --- | --- |
| Three-category* | 1 | 30,243 | 25.3 (0.844) | 7.1 (0.235) | 12 (0.400) | 21 (0.700) | 26 (0.867) | 28 (0.933) | 35 (1.167) | 76.4% |
|  | 2 | 21,268 | 85.2 (0.947) | 23.3 (0.259) | 59 (0.656) | 68 (0.756) | 82 (0.911) | 101 (1.122) | 129 (1.433) | 61.8% |
|  | 3 | 3,848 | 364.3 (0.998) | 69.5 (0.190) | 292 (0.800) | 327 (0.896) | 362 (0.992) | 399 (1.093) | 474 (1.299) | 51.6% |
| DPS<sup>†</sup> | 1 | 30,200 | 27.7 (0.923) | 7.4 (0.246) | 17 (0.567) | 24 (0.800) | 28 (0.933) | 33 (1.100) | 40 (1.333) | 61.4% |
|  | 2 | 21,156 | 84.7 (0.941) | 21.6 (0.240) | 49 (0.544) | 75 (0.833) | 84 (0.933) | 98 (1.089) | 117 (1.300) | 60.3% |
|  | 3 | 4,059 | 330.6 (0.906) | 64.5 (0.177) | 242.8 (0.665) | 304 (0.833) | 336 (0.921) | 367 (1.005) | 416 (1.140) | 74.3% |
| DPS, calibrated<sup>†</sup> | 1 | 30,155 | 29.4 (0.980) | 7.4 (0.246) | 17 (0.567) | 26 (0.867) | 31 (1.033) | 33 (1.100) | 40 (1.333) | 49.6% |
|  | 2 | 21,119 | 84.8 (0.942) | 20.2 (0.224) | 52 (0.578) | 75 (0.833) | 87 (0.967) | 96 (1.067) | 112 (1.244) | 60.4% |
|  | 3 | 4,153 | 316.2 (0.866) | 62.2 (0.170) | 234 (0.641) | 292 (0.800) | 322 (0.882) | 350 (0.959) | 395.8 (1.084) | 84.3% |

\* Reproduced from the baseline study [33] (Table 1). <sup>†</sup> Computed on the same 20 replications and simulation model as the baseline study; the uncalibrated DPS row reproduces the baseline DPS results.
Compliance = percentage of patients treated within MRWT. Proportions of MRWT shown in parentheses. Counts differ slightly across systems because re-prioritisation shifts which patients are treated within the analysis window.

The modest magnitude of this redistribution follows from the structure of the formula. The clinical factor enters priority only through the multiplier 1 + *c*, and calibration compresses the spread of *c*, its standard deviation falling from 0.19 to 0.10, whereas the time ratio *t*/*M* varies at least as widely and, unlike *c*, grows with every day a patient waits, so it dominates the ordering. The clinical term can therefore reorder only patients whose time ratios already lie close together, and the calibration, in compressing *c*, narrows that window further. This compression reflects the cross-procedure severity relationships elicited from the participating clinicians: a procedure-specific tool scales every procedure’s most severe state to the top of the unit interval, whereas the calibrated scale places those states at the levels the clinician consensus assigned, so the raw scores overstate the relative severity span of each procedure. The apparent dilution of the clinical term therefore arises because the formula was designed around scores carrying that artefact.

A control configuration that compressed the clinical-factor range identically but applied a single global band to every procedure, mapping each patient’s raw score linearly onto the pooled calibrated range [0.07, 0.70] irrespective of procedure, reproduced the compression while removing the cross-procedure re-ranking. This control accounted for most of the Category 3 reduction, a compression component averaging −10.4 days (95% confidence interval [−13.1, −7.8]), leaving a genuine cross-procedure re-ranking component of about −3 days (95% confidence interval [−5.5, −0.7]) that was small relative to the compression effect and inconsistent in direction, being absent or reversed in 8 of the 20 replications. The decomposition is, however, approximate, as the global band matches the span of the calibrated scores but not each procedure’s anchor positions, so the two components are not perfectly separated. The compression moves the system towards first-in-first-out admission, shortening the longest Category 3 waits and lengthening the shortest Category 1 waits irrespective of severity.

This preservation of severity concordance follows from a formula that weights the clinical term lightly, rather than from any shortcoming of the comparable scores; the urgency category already carries most of the severity ordering. The calibration’s value lies in the comparability of the scores, validated against clinician judgement (Kendall’s *τ* = −0.734); how heavily a prioritisation formula weights those comparable scores, rather than the calibration itself, governs their operational effect.

## 4 Discussion

This research has presented and demonstrated a structured calibration method designed to rescale procedure-specific PPTs onto a unified common scale. The strong alignment observed between calibrated scores and clinician judgements highlights the method’s internal consistency within this proof-of-concept study. This approach enables valid comparisons of clinical severity scores across different surgical procedures, promoting equitable prioritisation decisions. Such calibration addresses a practical challenge faced by hospitals worldwide: when patients awaiting different procedures compete for shared surgical resources, there is currently no principled basis for comparing their relative priority. The calibration method provides a transparent, clinician-informed mechanism for making these comparisons, thereby enabling both horizontal equity (similar clinical needs being similarly prioritised) and vertical equity (patients with greater needs being higher prioritised). Once calibrated, these priority scores can serve as coefficients in operations research scheduling models.

Overall, the operational analysis indicates that the value of calibration is realised at the level of the scores rather than of the existing formula. To examine this, the calibrated scores were substituted into the clinical-factor term of the dynamic priority score of Powers et al. [33], an existing prioritisation system evaluated through the same discrete-event simulation, changing only that term; embedded in this way, the calibrated scores redistributed waiting time between urgency categories while leaving cross-procedure severity concordance statistically unchanged. This outcome reflects the formula rather than the scale: the clinical term enters priority only through the multiplier 1 + *c*, which the dominant time ratio *t*/*M* outweighs, so that a more faithful severity signal cannot reorder patients whom the formula already separates by waiting time. The redistribution that does occur is therefore attributable mainly to the compression of the score range rather than to cross-procedure re-ranking. The Dynamic Priority Score is, in this respect, a conservative test of comparability, prioritising within a time-based frame in which the urgency category already carries most of the cross-procedure ordering. The setting that most requires a common scale is, by contrast, one in which patients awaiting different procedures compete directly for the same surgical resources, and such a setting has no principled basis for comparison without one. As such, calibration supplies a comparable severity signal that the current functional form weights too lightly to express, and the design of functional forms that draw more directly on this signal is taken up in future work.

For example, through the calibration method, a clinical factor score of 1 on the hernia PPT is equivalent to a score of 0.545 on the colectomy PPT, as can be seen in Table 2 (row 4). On the common scale, this is equivalent to a score of 0.439 (Table 3), as portrayed in Figure 10b. The clinical states from each of these PPTs corresponding to the aforementioned scores that can be considered “equal” can be seen in Figure 13. In the case of the colectomy PPT, the state presented is as close to a score of 0.545 as the weights allow (as described in Section 2.2.3).

**Figure 13:**
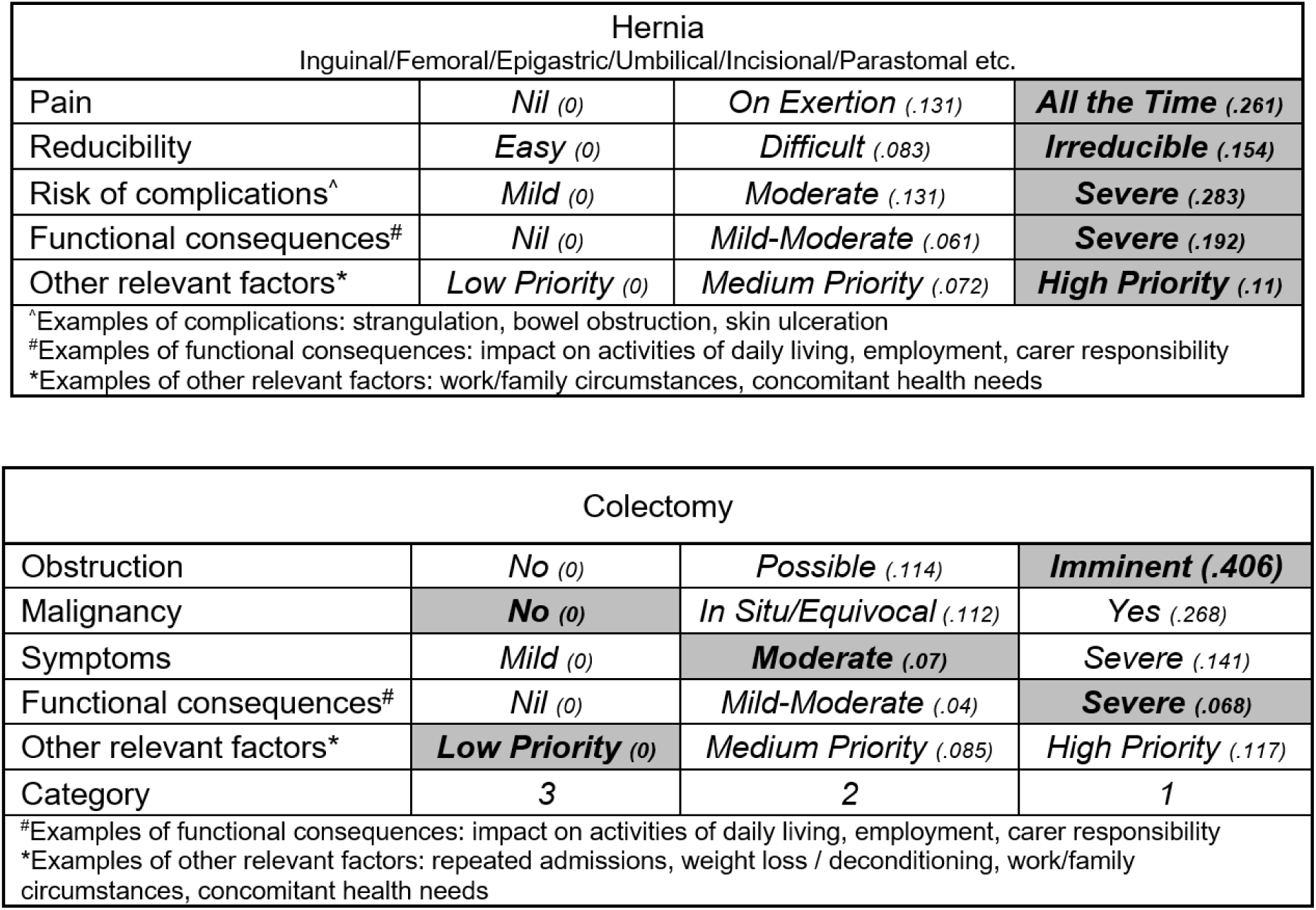
Hernia PPT with a clinical factor score of 1 and colectomy PPT with a clinical factor score of approximately 0.545, deemed equal on the common scale.

### 4.1 Limitations and Future Directions

This study represents a proof-of-concept demonstration conducted within a single institution. All PPTs, calibration exercises, and ranking assessments involved the same clinical team at one Australian public hospital, which ensured consistency in clinical judgement throughout but means that further validation with clinicians from other centres would be required to establish broader generalisability.

The operational analysis bounds the scope of these claims rather than the method itself: this study establishes that calibration yields comparable, clinically consistent scores, not that embedding them in an existing, time-ratio-dominated formula improves operational outcomes, since that depends on how heavily the formula weights the clinical term (Section 3.5). Realising the comparable signal operationally requires functional forms that weight it more heavily, a question beyond the scope of this study and taken up in future work.

A further consideration in applying the calibration method is the validity and reliability of existing PPTs. Calibrated scores rely directly on the appropriateness of the underlying criteria and weights, particularly for the base procedure, which in this study was colectomy. If clinician judgements are inconsistent with the PPT logic, inaccuracies could be propagated through the calibration process. Therefore, preliminary verification and refinement of PPTs is essential to ensure alignment with current clinical practices and expert evaluations.

Ideally, the same clinician group would collaboratively create and calibrate PPTs. However, practical constraints—including clinician availability, existing tool usage, and interdisciplinary requirements—often prevent this ideal scenario. Future applications of this calibration method should begin with a review of the underlying PPTs to confirm they remain consistent with current clinical practice. Clinicians should also be involved at an early stage in both the development and calibration phases to improve validity. When this is not feasible, routine validation through structured rankings and inter-rater reliability exercises using realistic or hypothetical patient scenarios can help mitigate these challenges [17].

The calibration method relies on linear scaling based on the assumption of interval-scale measurement properties for PPT scores, meaning score differences consistently reflect differences in clinical severity. Interval-scale measurement is also essential for tools used for cost-utility analysis or patient-reported outcome measures. Although this assumption generally holds true for PPTs developed using the PAPRIKA method and 1000minds software (as indicated by the software’s reported accuracy statistic, which improves when trade-off questions involve more than two criteria), it may not be universal.

For PPTs developed using other methods, interval-scale measurement may be inferred from the means of preference elicitation itself; otherwise, the method’s ability to produce interval-scale measurement should be tested in some way. When interval-scale measurement cannot be confidently assumed or attained, calibration may require additional equivalence points, leading to more complex transformation functions.

The use of extreme clinical states (most and least severe) as reference points in calibration may introduce inaccuracies if clinicians find these states unfamiliar or difficult to evaluate reliably. Although participants in this study found these extremes relatable and easily comparable, future research could benefit from incorporating more clinically familiar intermediate severity states to enhance reliability and clinician confidence.

Contradictions can highlight underlying mismatches between clinician intuition and the logical structure of the PPTs. Tracking the frequency and nature of contradictions can inform targeted refinements to PPTs, such as clarifying ambiguous criteria, simplifying terminology, or improving instructions and training for calibration participants. In future applications, a contradiction log could be used to identify patterns (e.g., which PPTs are most prone to inconsistent comparisons) and feedback could be collected immediately after a contradiction is registered. This feedback loop would enable iterative improvements to both the calibration process and the tools being calibrated, thereby increasing confidence in the comparability and fairness of prioritisation decisions.

Another potential consideration involves the cognitive load of the calibration exercise. Although the task of comparing patient states across procedures (Figure 6) may appear more demanding than traditional within-PPT trade-offs (Figure 4 and Figure 5), participants in this study reported that the process was straightforward when they were familiar with both procedures being compared.

Familiarity with the criteria and context of each PPT is critical to ensuring valid and confident judgements in pairwise comparisons.

Engaging a large number of clinicians from across a wide range of clinical backgrounds in the calibration method is also likely to improve accuracy. Where the calibration needs to span multiple specialties, each with a variety of PPTs, it would be useful to select a base PPT for each specialty, with inter-disciplinary calibration of pairs of base PPTs. This approach would allow further calibration of the specialty’s tools to be performed by clinicians with domain expertise. Additionally, the base PPTs could potentially be calibrated against a single general PPT to enable broader cross-specialty comparability.

In practice, the same calibration process applied in this study could be repeated within another surgical specialty to calibrate procedure-specific PPTs in that field. Once complete, those tools could then be integrated into the same common scale as the general surgery tools by calibrating one procedure from each specialty against a shared linking procedure. However, this approach may risk compounding errors if the linking procedures themselves are poorly calibrated or misaligned with clinical judgement. To address this, multiple linking procedures could be calibrated across specialties to provide cross-validation, improving the robustness and credibility of the resulting unified scale.

Furthermore, even within a single specialty, multiple base procedures could be used to calibrate other procedures in that discipline, providing internal validation and additional points of alignment to strengthen the common scale.

The calibration method can also be used in other types of health and non-health applications characterised by multiple decision tools with their own scales that need to be made comparable. For example, the decision-making and conjoint analysis platform 1000minds [37] has been used to create scoring tools across a wide range of sectors [36, 41]; see the brief surveys in the studies by Wijland et al. [42] and Sullivan et al. [43].

Among the most prominent collections of related but independent scoring tools created using 1000minds are those used to prioritise non-communicable [44], infectious [45] and antibiotic-resistant [46] diseases, respectively, for research purposes; using the calibration method, these three tools could be calibrated onto a common scale to produce an overall ranking of these diseases. Another example of a potential application is capital expenditure decision-making related to the provision of water services, where separate tools (like PPTs) could be used to prioritise capital investments in drinking water, wastewater, and stormwater infrastructure; a common scale could help decision-makers prioritise investments across all three water services together.

While our calibration method was specifically developed for aligning procedure-specific patient prioritisation tools, its flexibility suggests broader applicability. For example, the method could be applied to combine separate Best–Worst Scaling (BWS) utility scales derived from distinct healthcare decision contexts. One BWS study surveyed 214 patients across seven surgical oncology clinics about the factors influencing their choice of surgeon, quantifying trade-offs between clinical quality indicators (e.g., surgeon experience, hospital reputation) and access burdens (e.g., travel distance, waiting time) [47]. Another BWS study assessed public hospital outpatients’ preferences for appointment-related attributes, including diagnostic accuracy, continuity of care, wait times, and out-of-pocket costs [48]. While focused on different stages of care, these studies share overlapping criteria (particularly clinical quality and access burden), which provide anchor points for calibration.

However, the BWS utilities in each study exist on independent, arbitrary scales, and cannot be directly combined. To enable calibration, conceptually matched criteria must be identified, and multiple levels or approximations of attribute intensity may need to be defined to support iterative binary search comparisons. Provided these additional steps are implemented and interval-scale properties are confirmed, the resulting BWS utility scales could be calibrated onto a single joint utility function. This would enable policymakers to answer practical questions such as: *“How much additional waiting time or travel distance are patients willing to accept for higher-quality surgical care, and is this trade-off comparable to their willingness to wait for more accurate outpatient diagnosis?”* A unified scale of this kind could support more consistent, patient-centred service design, and help inform communication strategies that align with patient values, particularly in shared decision-making contexts where patients must weigh access constraints against the quality of care they receive.

Future research in the area of calibrating PPTs would expand the number of procedures and conditions made comparable on a common scale and investigate the extent to which calibration can adequately differentiate procedures within and between surgical specialties. As an initial pilot study, this research focused on a single specialty (general surgery) to demonstrate the feasibility of the approach.

Although the calibration method performed well in this context, future work is needed to explore its application across other specialties, incorporate a larger and more diverse sample of clinicians, and evaluate its performance on more complex or mixed sets of PPTs. An area of particular interest would be to apply the calibration method in a domain where both general and specific tools are concurrently used for the same patient. This would provide researchers with a unique opportunity to evaluate the accuracy and added value of the calibration method and, ultimately, reduce the burden of completing multiple PPTs for patients, clinicians, and administrators.

## 5 Conclusion

The calibration method developed and demonstrated in this study effectively transforms independently designed PPTs into a common severity scale. This unified scale provides a structured and transparent approach for comparing clinical severity across diverse procedures, supporting prioritisation decisions that reflect both horizontal equity (equal treatment of equal clinical need) and vertical equity (greater priority for greater clinical need). Given that hospitals routinely face the challenge of allocating shared surgical resources across patients awaiting fundamentally different procedures, this method offers a practical pathway towards more principled and defensible prioritisation.

The strong agreement observed between calibrated scores and clinician-derived rankings affirms the method’s validity and its capacity to reflect expert clinical judgement, and this agreement was maintained across the urgency categories of a full patient cohort. When placed within an existing dynamic priority formula, the calibrated scores redistributed waiting time between urgency categories rather than changing how well treatment order tracked severity, indicating that the operational value of the comparable signal depends on the functional form in which it is embedded. As this is a proof-of-concept study conducted within a single institution, further validation with clinicians from other centres would strengthen the generalisability of these findings.

In addition to demonstrating the method within a single specialty, this study lays the groundwork for broader applications. The approach can be adapted to other clinical specialties, and with appropriate validation, can support the integration of multiple tools across specialties into a single prioritisation framework. By using linking procedures and ensuring careful validation, the calibration method offers a scalable solution to the challenge of consistent patient prioritisation.

Furthermore, the method’s applicability is not limited to health care; it has potential in other fields where multiple independent tools or criteria must be aligned, such as resource allocation, capital investment prioritisation, and public policy planning.

Ultimately, this research represents a step towards fairer, more consistent prioritisation frameworks, and provides a foundation for future efforts to harmonise decision-making across disparate contexts using transparent, evidence-based calibration procedures.

## 6 Declarations

### 6.1 Ethics Approval and Consent to Participate

This study has ethics approval from The Prince Charles Hospital Human Research Ethics Committee (HREC/17/QPCH/336). Consent to participate not applicable.

### 6.2 Availability of Data and Materials

All data generated or analysed during this study are included in the tables herein. Any additional data referenced have been obtained from previously published sources, which have been appropriately cited. The calibration surveys are publicly accessible at the URLs listed in Appendix A and are provided under a CC BY 4.0 licence.

### 6.3 Competing Interests

FO and PH have ownership interests in 1000minds Ltd, whose software is used to calculate criterion weightings and implement the calibration method. All other authors declare no competing interests.

### 6.4 Funding

Financial support for this study was provided entirely through a SERTF grant from Sunshine Coast Hospital & Health Service, a government funded organisation (Queensland, Australia). The funding agreement ensured the authors’ independence in designing the study, interpreting the data, writing, and publishing the report. The following authors are employees of Sunshine Coast University Hospital: RA, SR, and DG. JP received a one-off student prize from 1000minds, including use of its software.

### 6.5 Authors’ Contributions

CRediT: JP: Conceptualisation, Data curation, Formal Analysis, Investigation, Methodology, Software, Validation, Visualisation, Writing – original draft, Writing – review & editing; FO: Methodology, Software, Writing – review & editing; PH: Methodology, Software, Writing – review & editing; RA: Conceptualisation, Data curation, Funding acquisition, Resources, Writing – review & editing; DG: Conceptualisation, Data curation, Funding acquisition, Writing – review & editing; SR: Conceptualisation, Data curation, Funding acquisition, Project administration, Resources, Writing – review & editing; JM: Conceptualisation, Supervision, Writing – review & editing; PC: Conceptualisation, Funding acquisition, Methodology, Project administration, Resources, Supervision, Writing – review & editing.

All authors read and approved the final manuscript.

## A Patient Prioritisation Tools and Calibration Survey Instruments

### Patient Prioritisation Tools (PPTs)

The 10 procedure-specific patient prioritisation tools (PPTs) used in this study were originally developed by clinicians from a public hospital in Australia. The full specifications of these PPTs (criteria, weights, and levels) are available from Figshare:

- Powers J, McGree JM, Grieve D, et al (2023) Managing surgical waiting lists through dynamic priority scoring – Datasets. figshare. https://doi.org/10.6084/m9.figshare. 19771738

These PPTs are licensed under a Creative Commons Attribution 4.0 International (CC BY 4.0) licence, permitting unrestricted reuse with proper attribution.

### Calibration Survey Instruments

Calibration was performed using structured online surveys in 1000minds. Each survey presented adaptive pairwise comparisons between patient states for two procedures, using the weightings developed in the aforementioned dataset.

The full calibration survey instruments are publicly available at the following permanent links:

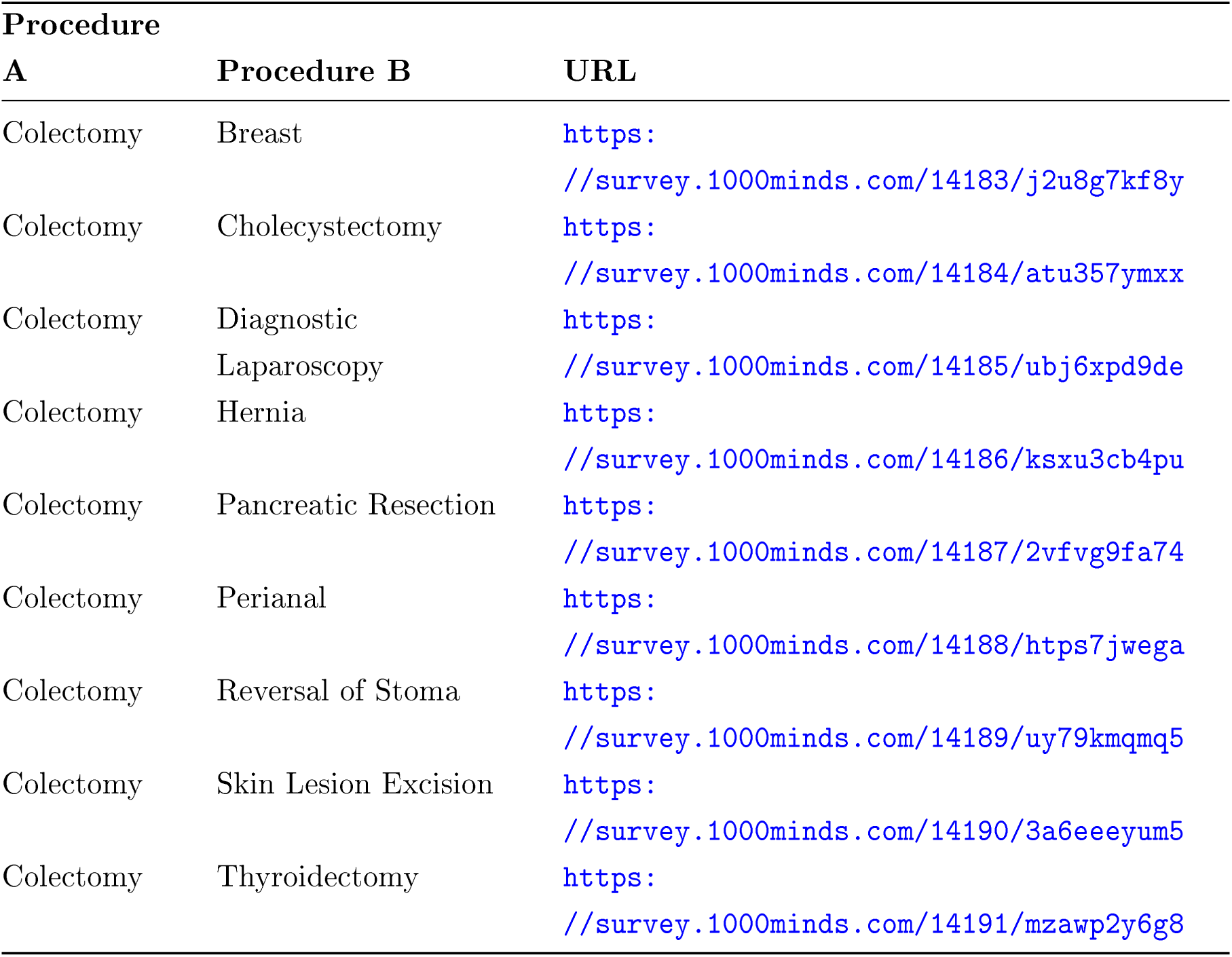

## Data Availability

All data produced in the present work are contained in the manuscript

## Notes

### Author Declarations

Ethics committee of The Prince Charles Hospital Human Research Ethics Committee (HREC/17/QPCH/336) gave ethical approval for this work.

